# The frequency of typhoid carriage in patients undergoing cholecystectomy for gallbladder disease in Pakistan: A cross-sectional study

**DOI:** 10.1101/2023.11.08.23298246

**Authors:** Sonia Qureshi, Noshi Maria, Tabish Chawla, Junaid Iqbal, Abdul Momin Kazi, Mehreen Adnan, Aneeta Hotwani, Najeeb Rahman, Muhammed Wahhaab Sadiq, Richelle Charles, Stephen Baker, Farah Naz Qamar

## Abstract

**Background:** Enteric fever is caused by *Salmonella* enterica serovars Typhi (*S*. Typhi) and Paratyphi A, B, and C. It continues to be a significant cause of morbidity and mortality worldwide. In highly endemic areas, children are disproportionately affected, and antimicrobial resistance reduces therapeutic options. It is estimated that 2–5% of enteric fever patients develop chronic asymptomatic infection. These carriers may act as reservoirs of infection; therefore, the prospective identification and treatment of carriers are critical for long-term disease control. We aimed to find the frequency of *Salmonella* Typhi carriers in patients undergoing cholecystectomy. We also compared the detection limit of culturing versus qPCR in detecting *S.* Typhi, performed a geospatial analysis of the carriers identified using this study, and evaluated the accuracy of anti-Vi and anti-YncE in identifying chronic typhoid carriage.

**Methods:** We performed a cross-sectional study in two centers in Pakistan. Gallbladder specimens were subjected to quantitative PCR (qPCR) and serum samples were analyzed for IgG against YncE and Vi by ELISA. We also mapped the residential location of those with a positive qPCR result.

**Findings:** Out of 988 participants, 3.4% had qPCR-positive gallbladder samples (23 *S.* Typhi and 11 *S.* Paratyphi). Gallstones were more likely to be qPCR positive than bile and gallbladder tissue. Anti-Vi and YncE were significantly correlated (r=0.78 p<0.0001) and elevated among carriers as compared to qPCR negative controls, except for anti-Vi response in Paratyphi A. But the discriminatory values of these antigens in identifying carriers from qPCR negative controls were low.

**Conclusion:** The high prevalence of typhoid carriers observed in this study suggests that further studies are required to gain information that will help in controlling future typhoid outbreaks in a superior manner than they are currently being managed.

**Author Summary:** Enteric fever, caused by bacteria *Salmonella* Typhi or Paratyphi, is a serious global illness particularly affecting children in areas with inadequate hygiene and sanitation facilities. It is transmitted through the oral-fecal route. In Pakistan, prevalence of extensively drug resistant *S.* Typhi resistant strain further complicates its treatment. Some infected individuals develop chronic infections and become carriers of these pathogens. These asymptomatic carriers may potentially spread the disease by shedding the bacteria in urine or stool. To assess the frequency of the carrier state, we examined gallbladder specimens (bile, stones, and tissue) from patients who underwent gallbladder removal surgeries at Aga Khan University and Jinnah Post-graduate Medical Centre. We demonstrated that PCR had a higher sensitivity in detecting *Salmonella* carriage with a detection limit of 10^2^ CFU/ml and identified 3.4% of typhoid carriers. Additionally, we investigated whether specific antibodies could identify these carriers using various serological tests. The results underscore the need for further research to promptly detect and appropriately treat this disease, thereby enhancing control and preventing future outbreaks.

## Introduction

Enteric fever, a collective term for typhoid and paratyphoid fevers, is caused by *Salmonella enterica serovars* Typhi (*S*. Typhi) and Paratyphi A, B, and C. It remains a major cause of morbidity and mortality worldwide, causing around 21 million new infections and 222,000 deaths annually (1). In highly endemic areas (Southeast Asia, South Central Asia, Latin America, and sub-Saharan Africa), children are disproportionately affected, and antimicrobial resistance reduces the therapeutic options available (2). In Pakistan, the annual typhoid incidence per 100,000 person-years is reported as 573.2 cases among 2-4 years and 412.9 cases among 5-15 years of age (3).

Enteric fever is acquired by ingestion of infecting microorganisms (4). After ingestion, the pathogens invade the intestinal epithelial barrier, where they are phagocytosed by macrophages, and then spread systematically to infect the liver, spleen, bone marrow, and gallbladder (5). It is estimated that 2–5% of enteric fever patients develop chronic asymptomatic infection (6). The chronic carriage can persist for years, and these individuals can intermittently shed the bacteria in stool or urine. It is estimated that ∼25% of asymptomatic chronic carriers have no clinical history of enteric fever(7, 8). These carriers may act as reservoirs of infection, therefore, the prospective identification and treatment of these carriers are critical for long-term disease control (9).

The detection of *Salmonella* carriage can be performed by isolating the organism from the stool, bile/duodenal contents, urine, gallbladder tissue or by microbiological techniques; serological methods; or molecular techniques (polymerase chain reaction (PCR) amplification). The sensitivity of a single stool culture for detecting carriers has been reported as 30-35%; therefore, multiple samples are required, which is logistically difficult at a population level. The sensitivity of a urine culture for detecting carriers is estimated to be less than 10% (10). *S*. Typhi has been isolated from the bile of patients undergoing cholecystectomy, which may be considered the gold standard for detection (11). However, this method may lack sensitivity in regions with excessive antimicrobial use (12, 13). Real-time quantitative PCR is a robust and sensitive method to detect and quantify bacteria in clinical samples and has been used to detect *S*. Typhi in blood, bone marrow, bile, and gallstones (14, 15). Additionally, the detection of IgG antibodies against Vi (anti-Vi) and YncE (anti-YncE) may provide an additional approach for diagnosing the carriage of invasive *Salmonella*.

Here, we aimed to measure the frequency and identify the associated factors of enteric fever carriers in patients undergoing cholecystectomy for gallbladder diseases in Pakistan by qPCR of bile, gallstones, and gallbladder tissue. An individual whose PCR was positive for *S.* Typhi or Paratyphi A in either of the specimens including bile, gallstone, or gallbladder tissue was considered a carrier. Moreover, we compared the detection limit of microbiology versus qPCR in detecting *Salmonella* organisms, performed a geospatial analysis of carriers identified using this study, and evaluated the accuracy of anti-Vi and anti-YncE in identifying chronic typhoid carriage.

## Methodology

### Study Design and Settings

A cross-sectional study was conducted at the Aga Khan University Hospital (AKUH) and Jinnah Postgraduate Medical Center (JPMC) from January 2019-March 2020. AKUH is a private tertiary care hospital based on ∼700 beds while JPMC is one of the largest government hospitals in Karachi city comprised of ∼1600 beds. This study was approved by the Ethical Review Committee (ERC) of both institutions.

### Patient Selection

Individuals of age ≥ 10 years who underwent cholecystectomy for gallbladder diseases were included in this analysis. Informed consent from participants/legal guardians and assent from children ≤ 18 years of age were obtained. Data was collected on a predesigned structured questionnaire that included information on participants’ demographics, medical history, and questions related to other independent factors like hand hygiene, history of typhoid fever, antibiotic use at the time of typhoid fever illness, overcrowding, etc.

### Gallbladder Specimen Collection and Transportation

Bile, gallbladder tissue, and gallstones were collected during cholecystectomy by the surgeons from their respective hospitals (AKUH and JPMC) in pre-labeled sterile tubes. Samples were transferred to Juma Research Laboratory (JRL) at the AKUH, Karachi maintaining 2-8^°^C. At JRL, all three specimens were stored at -80^°^C until further analysis.

### Assay for Determination of Limit of Detection of PCR

The pure culture of *S.* Typhi was grown overnight on Xylose Lysine Deoxycholate agar (XLD agar). A single colony was then inoculated in 4 mL of Luria Broth and incubated for 16 hours at 37°C to an optical density (OD) of 0.2 at 595 nm. This broth was then used to perform eight 10-fold serial dilutions in sterile Phosphate Buffer Saline (PBS). A 10µL aliquot of each dilution was plated in triplicate on XLD media. After 24 – 28hrs. of incubation at 37°C, plates with 30 to 300 colonies were counted. For each dilution, the average of the triplicate plates was calculated and the exact number of colony-forming units per milliliter was determined.

*S.* Typhi was also spiked 1:1 in sterile PBS and *S.* Typhi negative human bile sample. 900 µL of diluted bile was spiked with 100uL starting culture with an OD_595_ value of 0.2. 1mL of spiked bile sample was then 10-fold serially diluted fifteen times and then used for DNA extraction to perform qPCR.

### DNA extraction from gallbladder specimens

Bile, gallstone, and tissue samples once thawed at room temperature were subjected to DNA extraction. 25-30 mg of sample was used for extraction of DNA by Qiagen DNeasy blood & tissue kit (Qiagen, Hilden Germany) according to manufacturer’s protocol with modifications as noted below. For bile, 1 mL of each specimen was diluted with an equal volume of PBS, centrifuged at 13000 rpm for 10 minutes to concentrate bacterial cells in the pellet, the supernatant was discarded. Extracted DNA was stored at -80°C until qPCR was performed. For gallstone samples, the initial incubation with lysis buffer was optimized to 1 hour at 56^°^C.

### Primers and PCR Conditions

Real-time multiplex quantitative PCR was performed using primers and probes sequences as described by Nga et al, 2010. Primers and probes sequences were as follows; *S*. Typhi; ST-Frt 5’ CGCGAAGTCAGAGTCGACATAG 3’, ST-Rrt 5’ AAGACCTCAACGCCGATCAC 3’, ST-Probe 5’ FAM-CATTTGTTCTGGAGCAGGCTGACGG-TAMRA 3’ and S. Paratyphi A (SPA); Pa-Frt 5’ ACGATGATGACTGATTTATCGAAC 3’, Pa-Rrt 5’ TGAAAAGATATCTCTCAGAGCTGG 3’, SPA-Probe 5’ Cy5-CCCATACAATTTCATTCTTATTGAGAATGCGC-BBQ-650 3’(16).

qPCR reactions were carried out in 20 µL reaction volumes in a 96-well plate consisting of 2X TaqMan Fast Advanced Master mix and 5 µL of template DNA. Reaction concentrations of the two primer and probe sets for *S.* Typhi, and *S*. Paratyphi A were 0.4μM of each primer and 0.15μM of each probe. qPCR was performed on a Bio-Rad CFX96 real-time PCR system and fluorescence was released via the TaqMan 5’ to 3’ exonuclease activity. All qPCRs were cycled under the following conditions: 15 min at 95°C and 45 cycles of 30 Sec at 95°C, 30 Sec at 57.4°C and 30 Sec at 72°C. Water (no-DNA) was used as a negative template control (NTC). *S*. Typhi pure culture DNA was used as a positive control. A Ct value of ≤ 35 was considered positive.

### Laboratory assay for Vi and YncE IgG

An in-house kinetic ELISA was used to detect YncE antigen in serum as previously described (17). Briefly, 96-well plates were coated with YncE antigen (1μg/mL). Uncoated sites were blocked with 1% Bovine Serum Albumin (BSA). Following the washing phase, diluted serum samples (1:1000), blanks, and controls were added to the plate; controls included an internal typhoid-positive patient serum and a chimeric control (human anti-YncE IgG antibody). The Ag-Ab complex was subjected to goat anti-human IgG conjugated with horseradish peroxidase (Jackson Immuno Research), and peroxidase activity was measured at 405 nm using the ABTS substrate.

The concentration of anti Vi-IgG was measured using the VaccZyme Human anti-*S* Typhi Vi IgG Enzyme Immunoassay Kit (Binding site according to the manufacturer’s instructions. We used a standard curve to calculate the concentration of anti-Vi expressed in U/mL.

### Data Analysis

Statistical analysis was carried out using Stata software (version 12.0). Factor analysis was performed for constructing a wealth index for socioeconomic status. The crowding index was reported as ‘not crowded’ if the crowding index was ≤ 2 persons/room and considered as ‘crowded’ if the index was >2 persons/room. Cox proportional hazard algorithm was used to assess the association of typhoid carriers with independent variables. Multicollinearity among independent variables was assessed by using logistic commands. Results were reported as PR with 95% CI. p-values of ≤ 0.25 and ≤ 0.05 were considered significant for univariate and multivariable analysis respectively. Geographical location (GIS) mapping was done using Landsat satellite for visualizing the distribution of study participants. Patients residing in regions outside Karachi were identified and categorized according to the city. However, participants residing in Karachi were mapped according to their residential addresses. The relationship between anti-Vi and YncE responses was evaluated by spearman’s correlation coefficient and the Mann-Whitney U test was used to compare differences across groups. P≤0.05 were considered significant.

## Results

Out of 1,136 eligible patients, 1000 participants consented to participate and were enrolled. Statistical analysis was performed on the 988 participants with a gallbladder specimen, of which 668 participants (66.8%) had all three sample types: bile, gallstone, and gallbladder tissue (**Fig 1**). Of the 988 participants, 34 (3.4%) were detected as *Salmonella* carriers by qPCR. *S.* Typhi or Paratyphi detection was highest in PCR performed from gallstones specimens. (**Table 1**). Most of the typhoid carriers were ≤ 40 years of age (23/34, 67.6%) while the median (IQR) age in the typhoid carrier group was 37 (35–55) years compared to qPCR negative controls which had a median (IQR) of 45 (35–55) years. Most patients undergoing cholecystectomy were female, with no significant difference in percentage between *Salmonella* carriers and controls. Eight (23.5%) *Salmonella* carriers reported a history of having enteric fever or typhoid-like illness. The prevalence of a history of having enteric fever or typhoid-like illness was 5.75 times among the typhoid carriers compared to non-typhoid carriers after adjusting for other covariates. Most of the *Salmonella* carriers underwent cholecystectomy due to chronic cholecystitis with or without cholelithiasis (**Table 2**). The majority 30/34 (88.23%) of the *Salmonella* carriers were residents of Karachi and they were widely distributed all over the city (**Fig 2A, 2B and 2C**).

**Fig 1.**
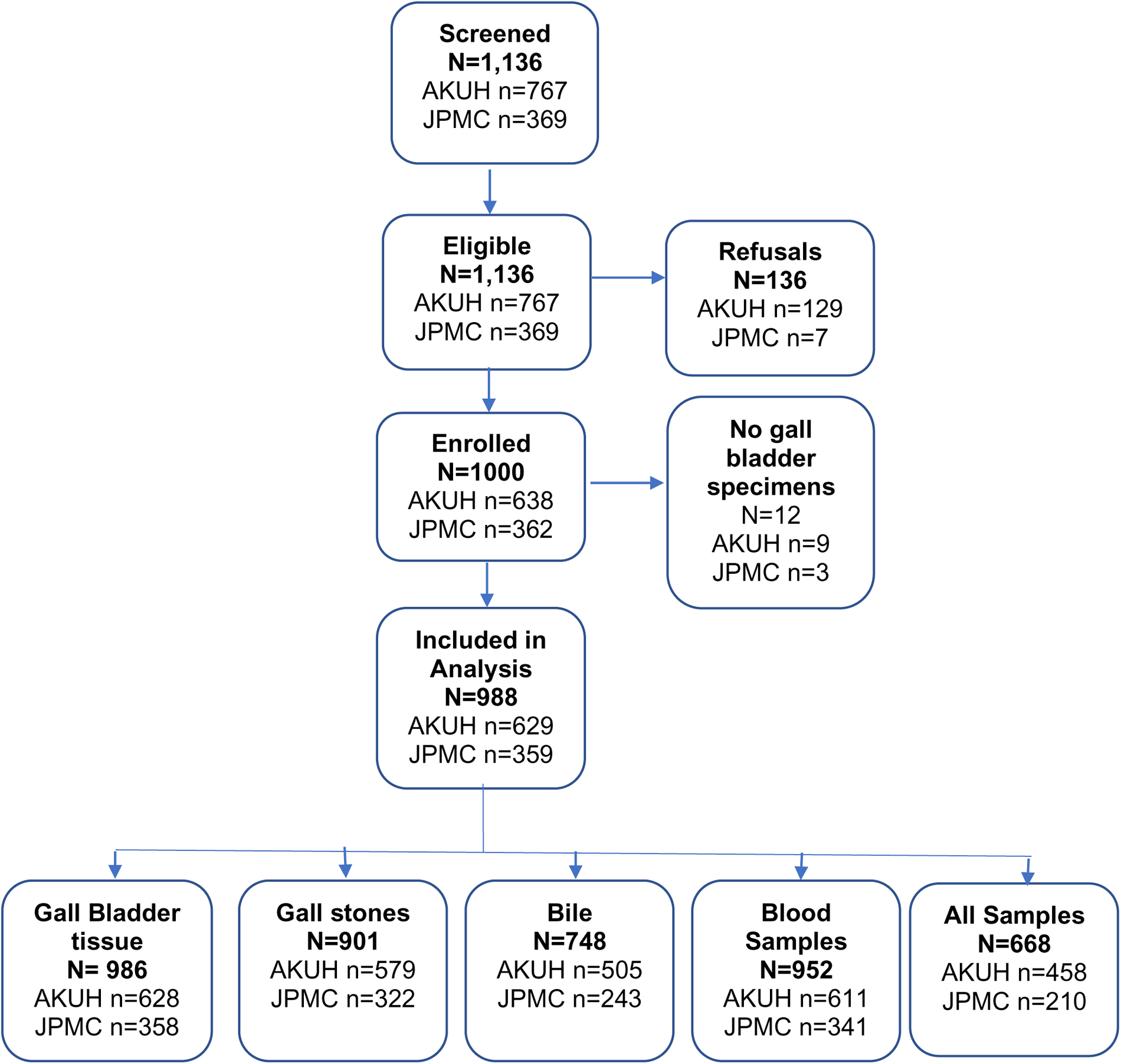
Study Flow.

**Fig 2A:**
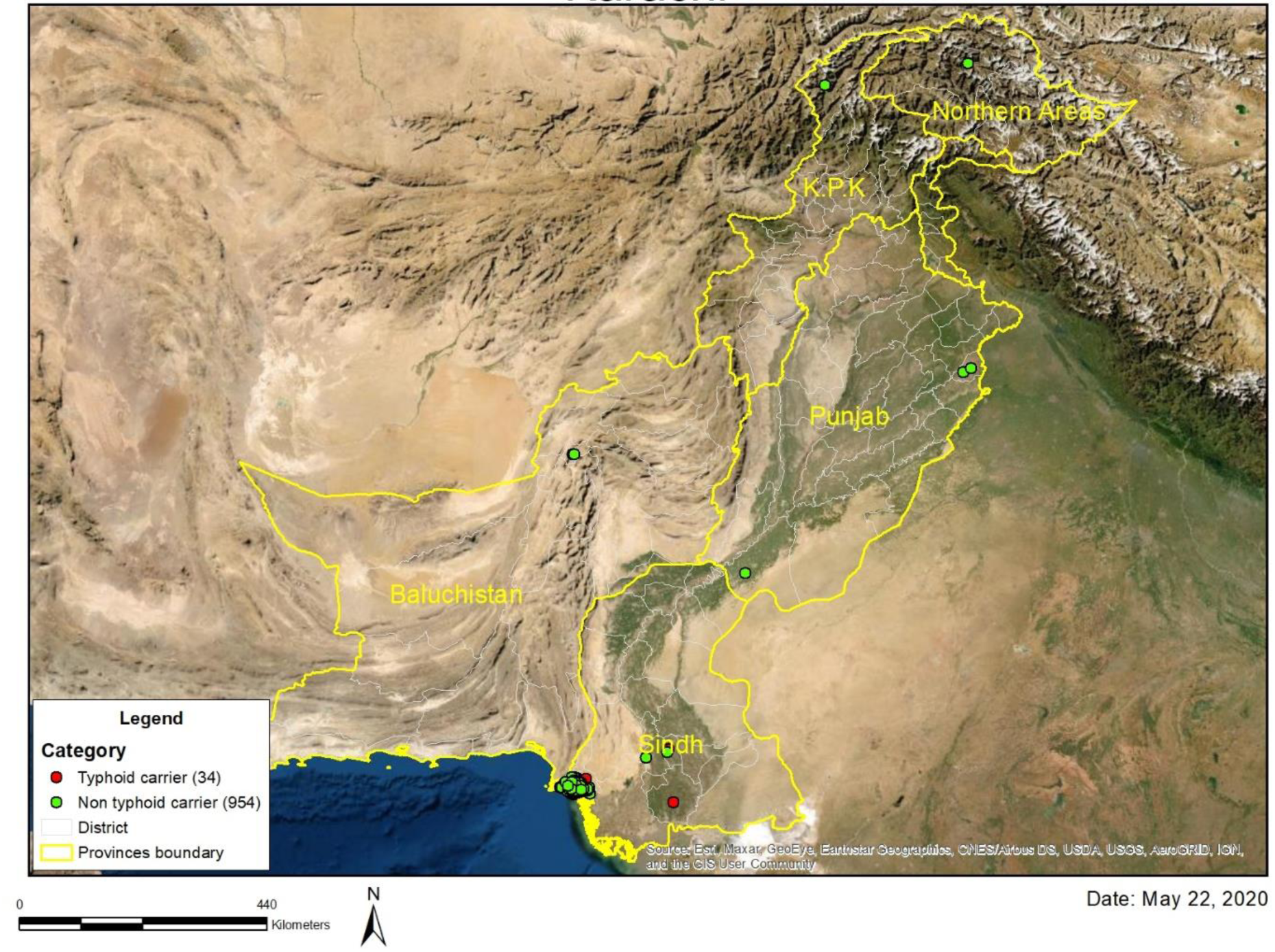
Geospatial map of Pakistan showing the distribution of study participants according to their geographical location as green (non-typhoid carriers) and red (typhoid carriers) dots.

**Fig 2B:**
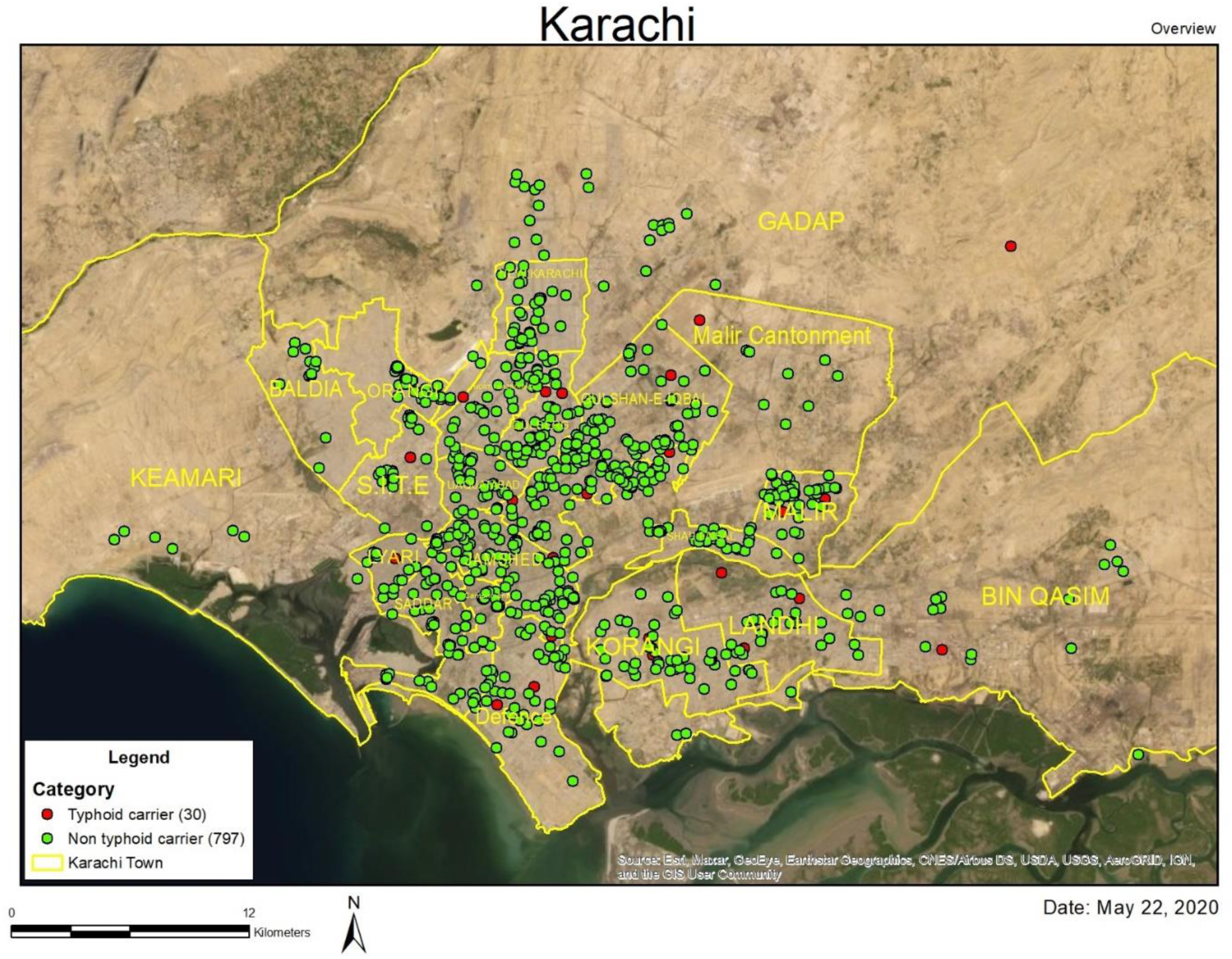
Geospatial map showing the distribution of typhoid carriers (red dots) and non-typhoid carriers (green dots) in Karachi, Pakistan.

**Fig 2C:**
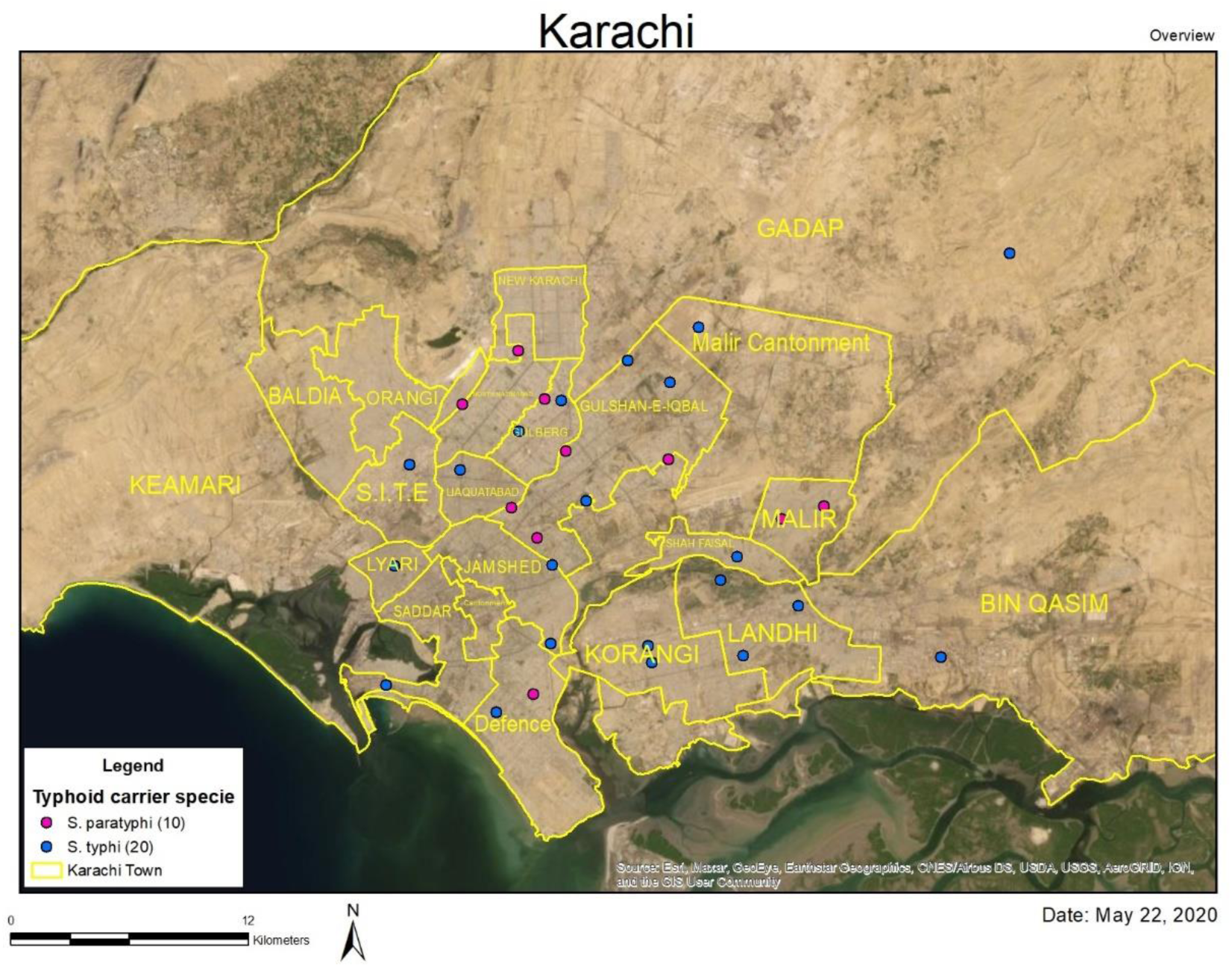
Geospatial map showing the distribution of typhoid carriers in Karachi Pakistan; blue dots (Salmonella Typhi) and pink dots (Salmonella Paratyphi A).

**Table 1:** PCR results for Salmonella Typhi and Paratyphi according to specimen type.

| S. No | Specimen Type | Salmonella Typhi | Salmonella Paratyphi | Total |
| --- | --- | --- | --- | --- |
|  |  | n/N (%) | n/N (%) | n/N (%) |
|  | Aga Khan University Hospital (AKUH) |  |  |  |
| 1 | Bile | 2/505 (0.40) | 0/505 (0.00) | 2/505 (0.40) |
| 2 | Gall stone | 5/579 (0.86) | 1/579 (0.17) | 6/579 (1.04) |
| 3 | Gall bladder tissue | 1/628 (0.16) | 2/628 (0.32) | 3/628 (0.48) |
| 4 | Stone and Tissue | 1/469 (0.21) | 1/469 (0.21) | 2/458 (0.44) |
| 5 | Stone, Tissue, and Bile | 2/469 (0.21) | 1/469 (0.21) | 3/469 (0.64) |
|  | Total | 11/630 (1.75) | 5/630 (0.79) | 16/630 (2.54) |
|  | Jinnah Postgraduate Medical Center (JPMC) |  |  |  |
| 1 | Bile | 0/243 (0.00) | 0/243 (0.00) | 0/243 (0.00) |
| 2 | Gall stone | 1/322 (0.31) | 3/322 (0.93) | 4/322 (1.24) |
| 3 | Gall bladder tissue | 4/358 (1.11) | 0/358 (0.00) | 4/358 (1.11) |
| 4 | Stone and Tissue | 3/210 (1.42) | 1/210 (0.48) | 4/210 (1.90) |
| 5 | Bile and Tissue | 1/210 (0.48) | 0/210 (0.00) | 1/210 (0.48) |
| 6 | Bile and Stone | 1/210 (0.48) | 1/210 (0.48) | 2/210 (0.95) |
| 7 | Stone, Tissue, and Bile | 2/210 (0.95) | 1/210 (0.48) | 3/210 (1.42) |
|  | **Total** | **12/358 (3.35)** | **6/358 (1.68)** | **18/358 (5.02)** |

**Table 2:**
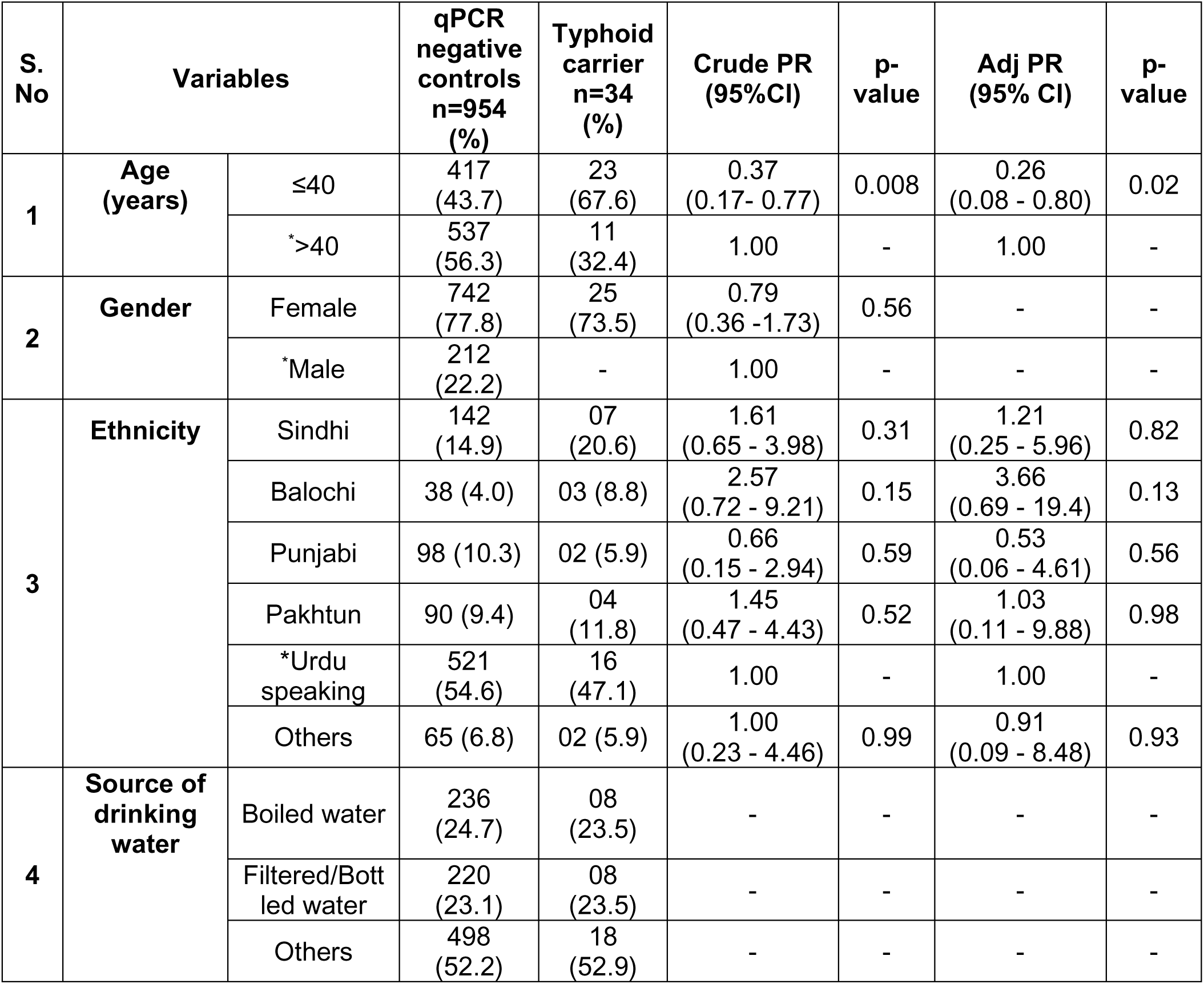
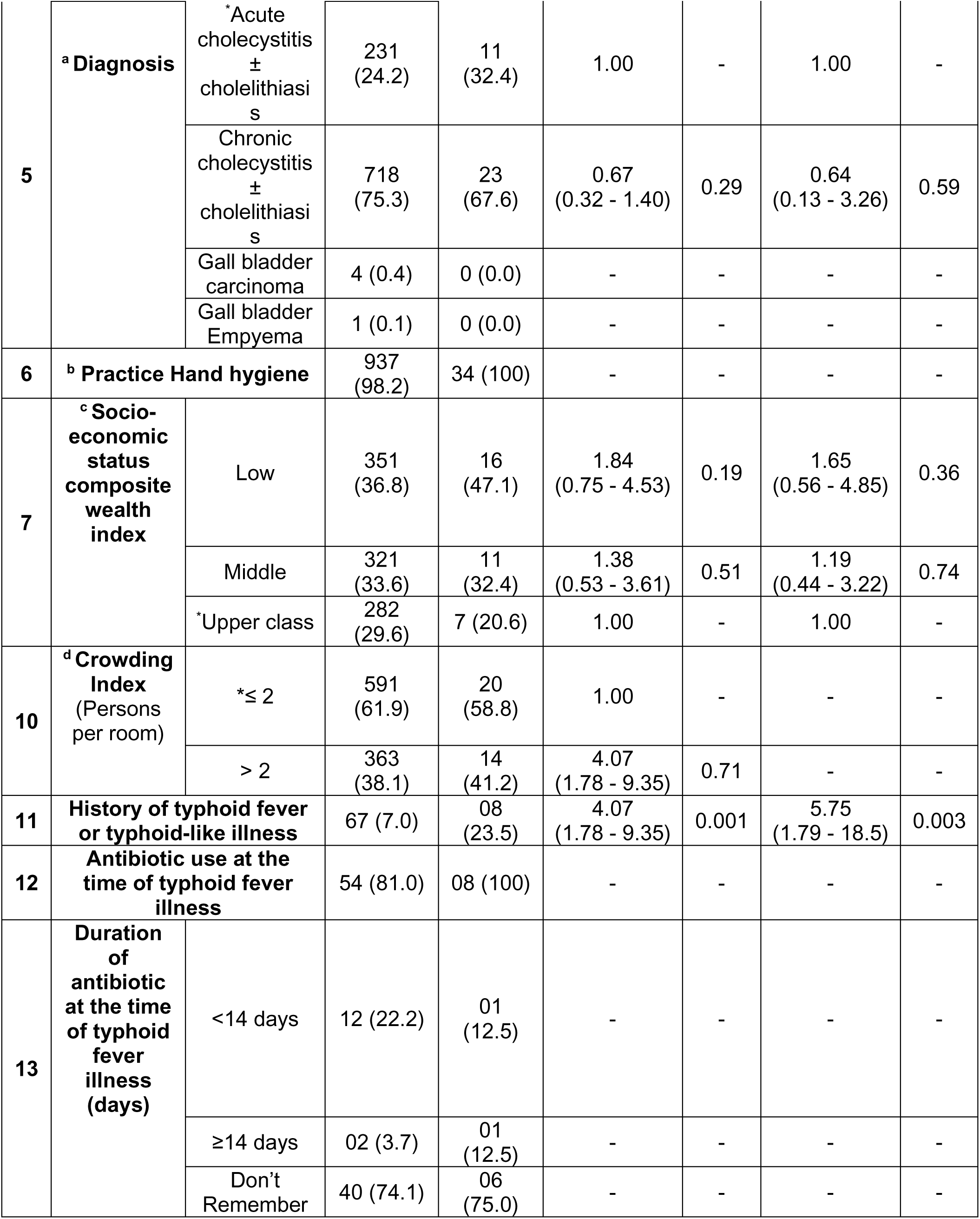
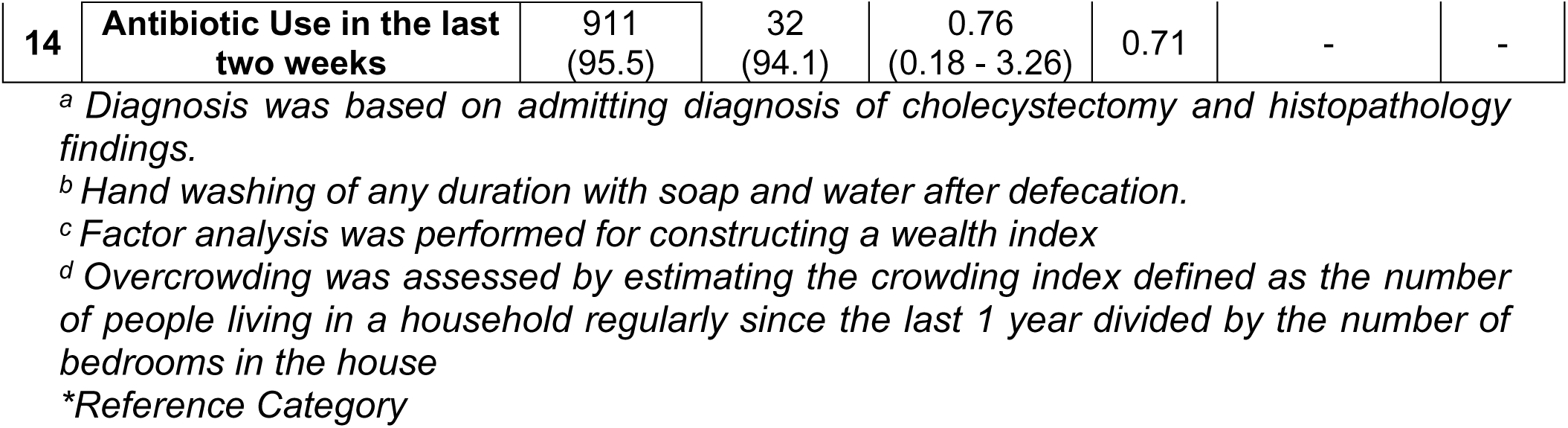
Descriptive statistics, unadjusted and adjusted analysis for various factors for typhoid carriers among patients who underwent cholecystectomy for gall bladder diseases.

| S. No | Variables |  | qPCR negative controls n=954 (%) | Typhoid carrier n=34 (%) | Crude PR (95%CI) | p-value | Adj PR (95% CI) | p-value |
| --- | --- | --- | --- | --- | --- | --- | --- | --- |
| 1 | Age (years) | ≤40 | 417 (43.7) | 23 (67.6) | 0.37 (0.17- 0.77) | 0.008 | 0.26 (0.08 - 0.80) | 0.02 |
|  |  | * >40 | 537 (56.3) | 11 (32.4) | 1.00 | - | 1.00 | - |
| 2 | Gender | Female | 742 (77.8) | 25 (73.5) | 0.79 (0.36 -1.73) | 0.56 | - | - |
|  |  | *Male | 212 (22.2) | - | 1.00 | - | - | - |
| 3 | Ethnicity | Sindhi | 142 (14.9) | 07 (20.6) | 1.61 (0.65 - 3.98) | 0.31 | 1.21 (0.25 - 5.96) | 0.82 |
|  |  | Balochi | 38 (4.0) | 03 (8.8) | 2.57 (0.72 - 9.21) | 0.15 | 3.66 (0.69 - 19.4) | 0.13 |
|  |  | Punjabi | 98 (10.3) | 02 (5.9) | 0.66 (0.15 - 2.94) | 0.59 | 0.53 (0.06 - 4.61) | 0.56 |
|  |  | Pakhtun | 90 (9.4) | 04 (11.8) | 1.45 (0.47 - 4.43) | 0.52 | 1.03 (0.11 - 9.88) | 0.98 |
|  |  | *Urdu speaking | 521 (54.6) | 16 (47.1) | 1.00 | - | 1.00 | - |
|  |  | Others | 65 (6.8) | 02 (5.9) | 1.00 (0.23 - 4.46) | 0.99 | 0.91 (0.09 - 8.48) | 0.93 |
| 4 | Source of drinking water | Boiled water | 236 (24.7) | 08 (23.5) | - | - | - | - |
|  |  | Filtered/Bott led water | 220 (23.1) | 08 (23.5) | - | - | - | - |
|  |  | Others | 498 (52.2) | 18 (52.9) | - | - | - | - |
| 5 | <sup>a</sup>Diagnosis | *Acute cholecystitis ± cholelithiasis | 231 (24.2) | 11 (32.4) | 1.00 | - | 1.00 | - |
|  |  | Chronic cholecystitis ± cholelithiasis | 718 (75.3) | 23 (67.6) | 0.67 (0.32 - 1.40) | 0.29 | 0.64 (0.13 - 3.26) | 0.59 |
|  |  | Gall bladder carcinoma | 4 (0.4) | 0 (0.0) | - | - | - | - |
|  |  | Gall bladder Empyema | 1 (0.1) | 0 (0.0) | - | - | - | - |
| 6 | <sup>b</sup> Practice Hand hygiene |  | 937 (98.2) | 34 (100) | - | - | - | - |
| 7 | <sup>c</sup> Socio-economic status composite wealth index | Low | 351 (36.8) | 16 (47.1) | 1.84 (0.75 - 4.53) | 0.19 | 1.65 (0.56 - 4.85) | 0.36 |
|  |  | Middle | 321 (33.6) | 11 (32.4) | 1.38 (0.53 - 3.61) | 0.51 | 1.19 (0.44 - 3.22) | 0.74 |
|  |  | *Upper class | 282 (29.6) | 7 (20.6) | 1.00 | - | 1.00 | - |
| 10 | <sup>d</sup> Crowding Index (Persons per room) | *≤ 2 | 591 (61.9) | 20 (58.8) | 1.00 | - | - | - |
|  |  | > 2 | 363 (38.1) | 14 (41.2) | 4.07 (1.78 - 9.35) | 0.71 | - | - |
| 11 | History of typhoid fever or typhoid-like illness |  | 67 (7.0) | 08 (23.5) | 4.07 (1.78 - 9.35) | 0.001 | 5.75 (1.79 - 18.5) | 0.003 |
| 12 | Antibiotic use at the time of typhoid fever illness |  | 54 (81.0) | 08 (100) | - | - | - | - |
| 13 | Duration of antibiotic at the time of typhoid fever illness (days) | <14 days | 12 (22.2) | 01 (12.5) | - | - | - | - |
|  |  | ≥14 days | 02 (3.7) | 01 (12.5) | - | - | - | - |
|  |  | Don't Remember | 40 (74.1) | 06 (75.0) | - | - | - | - |

|  |  |  |  |  |  |  |  |
| --- | --- | --- | --- | --- | --- | --- | --- |
| 14 | Antibiotic Use in the last two weeks | 911<br>(95.5) | 32<br>(94.1) | 0.76<br>(0.18 - 3.26) | 0.71 | - | - |

<sup>a</sup> Diagnosis was based on admitting diagnosis of cholecystectomy and histopathology findings.
<sup>b</sup> Hand washing of any duration with soap and water after defecation.
<sup>c</sup> Factor analysis was performed for constructing a wealth index
<sup>d</sup> Overcrowding was assessed by estimating the crowding index defined as the number of people living in a household regularly since the last 1 year divided by the number of bedrooms in the house
\*Reference Category

### YncE and Vi antibody responses

YncE IgG ELISAs were performed on all patients with available serum (33 carriers, 919 PCR negative controls) and Vi IgG was measured in 33 carriers and 32 PCR negative age (± 01 years) and gender matched controls. A positive correlation was observed between serum Vi and YncE antibody responses among carriers (r=0.78, p<0.0001), and the correlation was highest in patients that were PCR positive. This was observed from multiple sites (**Fig 3A**). This correlation was weaker when including the full cohort of qPCR negative and positive patients (r=0.55, p<0.0001) (**Fig 3A**). YncE and Vi values were significantly higher in PCR-positive patients compared to PCR-negative controls (**Fig 3B**), except for anti-Vi levels in *S*. Paratyphi A carriers where no difference was seen between groups.

**Fig 3.**
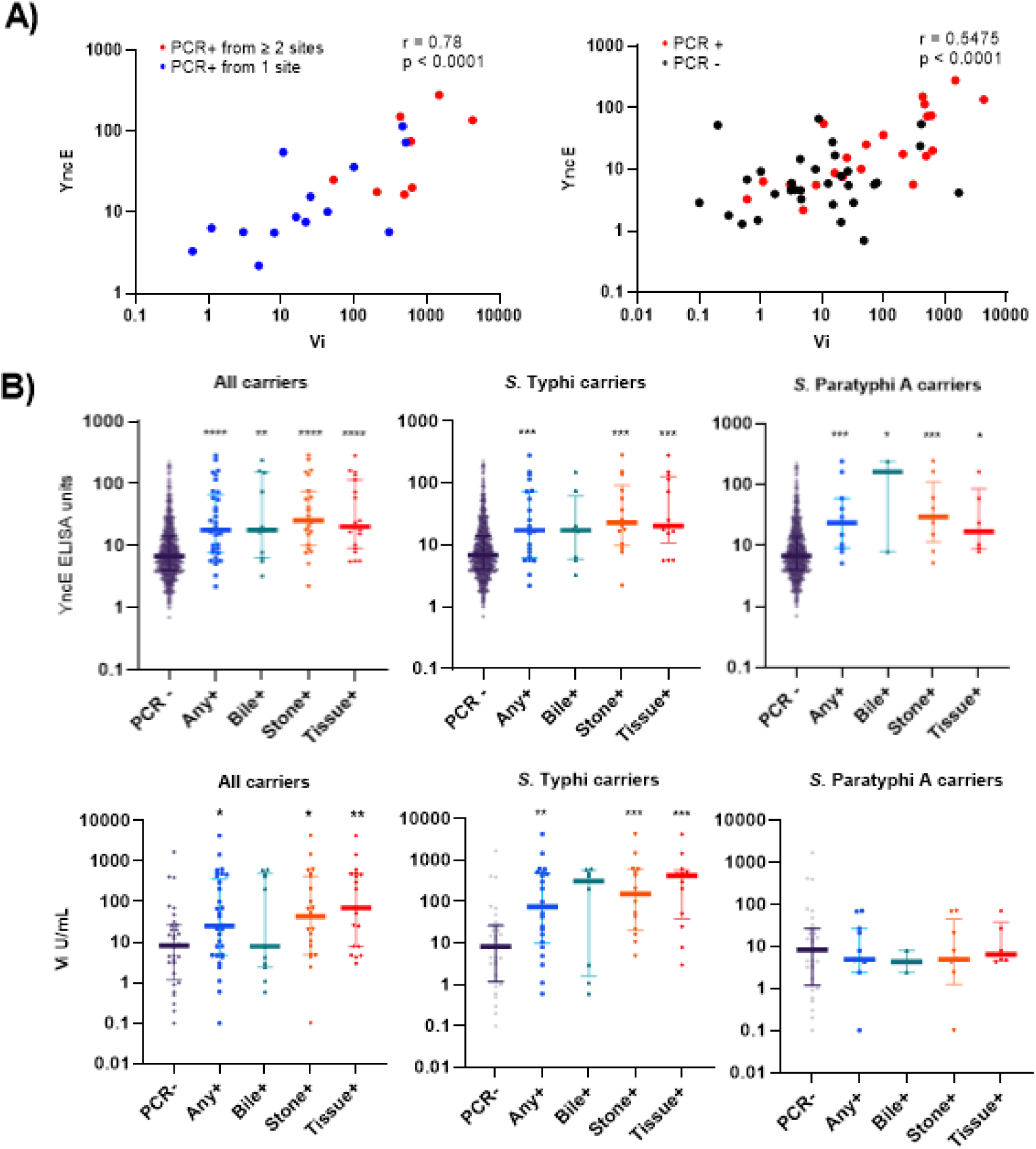
Anti-YncE and Vi IgG levels in *S.* Typhi or *S.* Paratyphi A qPCR positive and negative individuals. (A) Spearman correlation (B) Distribution of anti YncE and Vi levels by PCR status and site of positivity. Data are shown as median with IQR. Statistical analysis by Mann-Whitney U test. * p<0.05; ** p<0.01 ***p<0.001; ****p<0.0001

We assessed the classification accuracy of each antigen using the receiver operating characteristic (ROC) area under the curve (AUC). We found that both antigens had relatively poor accuracy in distinguishing PCR-positive carriers from PCR-negative controls underwent cholecystectomy (**Table 3**). The AUC for YncE for the following carrier cohorts compared to PCR negative controls was 0.73 for all carriers, 0.70 for Typhi carriers, and 0.78 for Paratyphi A carriers. For Vi, the AUC was 0.66 for all carriers, 0.75 for Typhi carriers, and 0.52 for Paratyphi A carriers. The discrimination accuracy for YncE and Vi did differ by the type of sample that was PCR positive. For *S*. Paratyphi A carriers, PCR positive by bile and stone with an AUC of 0.85 and 0.81, respectively for YncE and the discriminatory accuracy of Vi was highest for *S.* Typhi carriers, PCR positive by stone or tissue, AUC 0.83 and 0.84, respectively. This sub-analysis, however, was limited by low numbers.

**Table 3:** Accuracy of distinguishing Typhi carriers (*S*. Typhi or *S*. Paratyphi A) from qPCR negative controls using anti-YncE and Vi IgG responses.

|  |  | All carriers | *S. Typhi* carriers | *S. Paratyphi A* carriers |
| --- | --- | --- | --- | --- |
| Antigen | Site | AUC (95% CI) | AUC (95% CI) | AUC (95% CI) |
| YncE | Any | 0.73 (0.65 - 0.82) | 0.70 (0.59 - 0.83) | 0.78 (0.67 - 0.90) |
| Vi |  | 0.66 (0.53 - 0.79) | 0.75 (0.61 - 0.88) | 0.52 (0.32 - 0.71) |
| YncE | Bile | 0.73 (0.57 - 0.88) | 0.68 (0.49 - 0.87) | 0.85 (0.62 - 1.00) |
| Vi |  | 0.60 (0.39 - 0.82) | 0.69 (0.43 - 0.94) | 0.61 (0.43 - 0.80) |
| YncE | Stone | 0.79 (0.70 - 0.89) | 0.78 (0.65 - 0.91) | 0.81 (0.68 - 0.95) |
| Vi |  | 0.68 (0.54 - 0.83) | 0.83 (0.71 - 0.95) | 0.55 (0.33 - 0.76) |
| YncE | Tissue | 0.78 (0.69 - 0.88) | 0.79 (0.67 - 0.91) | 0.77 (0.64 - 0.91) |
| Vi |  | 0.76 (0.62 - 0.89) | 0.84 (0.71 - 0.98) | 0.57 (0.37 - 0.77) |

### Analytical sensitivity of bile qPCR

The detection limit of qPCR using bile samples was determined using bile samples spiked with decreasing 10-fold concentrations *of in-vitro-grown S.* Typhi (**Fig 4**). The limit of detection of the assay was found to be 16 cells.

**Fig 4:**
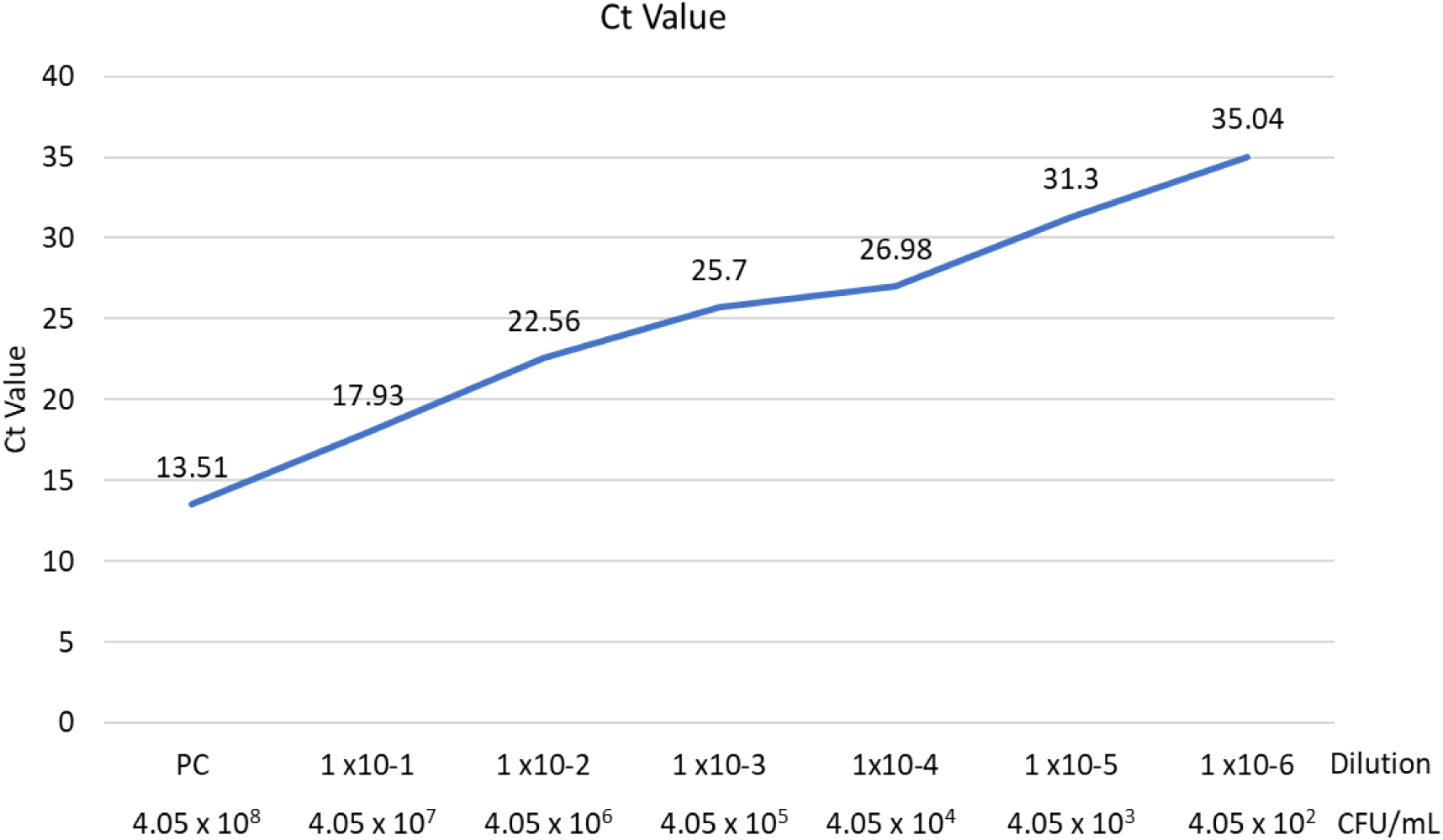
Cycle Quantification (Cq) values and colony forming unit (CFU/ml) count at different dilutions of *S*. Typhi in Bile.

## Discussion

Currently, there are no available methods to predict who will become a carrier after acute infection (18). Recovery from the acute phase of the illness is possible with appropriate antimicrobial treatment, however, a small fraction of individuals develop persistent asymptomatic infection (19) and may serve as the source of outbreaks in endemic communities.

In this study, we demonstrated that PCR had a higher sensitivity to detect Salmonella carriage with a detection limit of 10^2^ CFU/ml. Of the 988 enrolled participants, we identified 34 (3.4%) typhoid carriers by PCR. This prevalence is in line with the results of studies conducted in other endemic regions i.e., 2-5 % (20, 21). Other PCR-based studies from the high endemic region have reported a higher prevalence 17.5% and 10.1% with zero and one culture positivity, respectively (22, 23). These findings confirm the presence of nonculturable *S.* Typhi and *S.* Paratyphi persisting in the gallbladder, which may be due to prior antibiotic exposure, presence in a viable but nonculturable state in the biliary system; or they may be present in sparse numbers and thus sensitive to sampling bias (24).

There were higher anti-Vi and YncE responses between carriers and qPCR negative controls, with a significant positive correlation between these antibody responses amongst carriers. Anti-Vi, however, was not elevated in *S*. Paratyphi A carriers as this organism does not express Vi. These findings are in line with prior studies that showed promise for these antigens to detect typhoid carriers (17, 24–27). However, the diagnostic accuracy of anti-Vi IgG and YncE IgG to distinguish PCR-positive typhoid biliary carriers from PCR-negative individuals undergoing elective cholecystectomy was low. The performance of anti-Vi was comparable to the prior study of bile culture-confirmed patients underwent elective cholecystectomy, but YncE IgG did not perform well in this cohort (26, 28). PCR-positive, culture-negative carriers may have a lower burden of disease, leading to lower-level antibody responses. We did see higher antibody positivity in patients that were positive by multiple sample types which would support this hypothesis. The limited specificity observed for these antigens could be due to various factors, including potential misclassification of carriers as PCR negative due to the assay’s detection limit. Moreover, other factors beyond carriage, such as recent infections or cross-reactivity with other organisms carrying similar genes (i.e. *Citrobacter* for Vi or *E. coli* for YncE) could contribute to elevated Vi and YncE responses among PCR negative carriers. It is also possible that the PCR-positive individuals may represent recently infected individuals, rather than chronic carriers since this was a cross-sectional analysis. A large community-based study in Vietnam showed a prevalence of elevated Vi responses of 3%, however, *S*. Typhi was never detected in stool. This study suggests that in endemic zones, Vi responses may be detected in both chronic carriers and recently infected individuals (28). YncE is expressed in a small percentage of acutely infected individuals as well. Thus, the utility for identifying carriers using available serological techniques, including both YncE and Vi, in general endemic populations and even in this higher-risk cohort, remains unclear. These antigens may still be useful in outbreak studies for the detection of chronic carriers when a small number of participants are implicated (e.g., food handlers).

Like prior studies, our data demonstrates that the majority of carriers are female (>70%) (6, 18), Since there was no difference between the percentage of female *Salmonella* carriers and non-carriers, this association is likely due to the increased incidence of cholecystitis and cholelithiasis among women (29, 30). 23.5% of patients that were diagnosed as *Salmonella* carriers reported a history of enteric fever or enteric fever-like illness. This low prevalence might be due to recall bias, getting antibiotic prescriptions without laboratory diagnosis (only 61 participants reported that they did the blood culture investigation when they acquired typhoid fever in past and none could recall a positive culture), or subclinical infections. Despite this, having a history of enteric fever or enteric fever-like illness was 5.75 times higher among *Salmonella* carriers compared to non-*Salmonella* carriers after adjusting for other covariates.

A limitation of our analysis is that we have an overrepresentation in our cohort of patients recruited from The Aga Khan university hospital (AKUH), a private hospital, compared to Jinnah Postgraduate Medical Centre (JPMC), a government hospital. The socioeconomic status of the populations served in these two tertiary care hospitals differs significantly. Despite this, we were able to detect a difference in the prevalence of carriers, which was higher in JPMC (5.02%) compared to AKUH (2.54%). This reflects the need for future studies in low socioeconomic regions to obtain a better estimate of the true prevalence of *Salmonella* carriage and associated risk factors.

## Conclusion

The current study has demonstrated the significant prevalence of biliary *S*. Typhi and Paratyphi carriage in Karachi, Pakistan which is alarming for further outbreaks. Although anti-Vi and YncE IgG are elevated in typhoid carriers, the diagnostic accuracy of these antigens in detecting *Salmonella* carriage in a general endemic population undergoing elective cholecystectomy was low. Improved strategies for simple, rapid identification of carriers within a community for timely identification and appropriate treatment of this preventable risk factor can play a significant role in typhoid amelioration.

## Data Availability

Data will be available upon request.

## Author Contributions

This study was conceptualized by SQ, SB, and FNQ. Contribution in design of the study was done by SQ, JI, RC, SB, and FNQ. Data collection and data management were handled by SQ, TC, NM, MA, AH, and NR. Data analyses and interpretation were carried out by SQ, NR, NM, JI, AMK, RC, SB, and FNQ. The manuscript was drafted by SQ and NM with input from MA, AH, RC, MWS and FNQ. All the authors reviewed manuscript drafts, provided inputs, and approved the final version.

## Acknowledgments

We are grateful to the surgeons, surgery trainees, staff members, technicians, and nurses of AKUH and JPMC for providing gallbladder specimens for this research. We are thankful to our co-workers, data collectors, data entry operators, and data management team for showing their dedication and determination throughout this study.

## Ethical Approval

This study was approved by the Ethical Review Committees (ERC) of Aga Khan University Hospital and Jinnah Postgraduate Medical Center.

